# A population-based seroprevalence survey of severe acute respiratory syndrome coronavirus 2 infection in Beijing, China

**DOI:** 10.1101/2020.09.23.20197756

**Authors:** Xiaoli Wang, Wenjing Gao, Shujuan Cui, Yi Zhang, Ke Zheng, Ji Ke, Jun Lv, Canqing Yu, Dianjianyi Sun, Quanyi Wang, Liming Li

## Abstract

**BACKGOUND:** The spread of Coronavirus Disease 2019 (COVID-19) had been controlled in China. The seroprevalence of COVID-19 in Beijing has not been evaluated.

**METHODS:** In April, residents in Beijing were randomly enrolled. Blood samples were collected and antibodies to SARS- CoV-2 were tested by two colloidal gold kits. All colloidal gold positive serums were then tested by Micro- neutralization assay.

**RESULTS:** None of 2,184 residents participated was tested positive by micro-neutralization assay. The seroprevalence of COVID-19 in Beijing was estimated < 0.17%.

**CONCLUSIONS:** The seroprevalence of COVID-19 was low in April suggesting that community-wide spread was prevented in Beijing.

## Introduction

Coronavirus Disease 2019 (COVID-19) has rapidly spread throughout the world. Reported cases only represented a small fraction of the real number of cases, since most of asymptomatic cases were hardly detected. According to a few early studies from Iceland, US and Iran, the seropositivity at community level ranged from 0.6%-22%, much higher than the level of reported incidence rate [1-4]. The rate of undetected infections also varied in different countries. Investigating seroprevalence in different locations around the world enables further understanding the infection spectrum, the incidence and case-fatality rate, and most importantly, population immunity level of syndrome coronavirus 2 (SARS-CoV-2). Seroprevalence is also a key factor to optimize strategies for disease prevention and control. Serological surveys enable the determination of infection susceptibility, past acute infection and recovery group that has potentiality immune to re-infection. Thus, it is essential to conduct a large-scale serological survey to SARS-CoV-2.

COVID-19 was first reported in Wuhan, China. With the rapid spread of SARS-CoV-2 all around the world, Beijing - the political, cultural and international exchange center of China- faced an extraordinary challenge both at home and abroad. The first COVID-19 case in Beijing was confirmed on January 19^th^, 2020. The authority in Beijing implemented a series control measures to prevent further spread during the past several months. As of April 15, a total of 594 COVID-19 cases including 9 deaths were officially confirmed and reported in Beijing, with an incidence rate of 2.75/100,000 and a case-fatality rate of 1.5%.

To evaluate the effectiveness of control strategies and the risk of future epidemic in Beijing, a community-based serological survey was performed.

## Method

### Study design

A community-based, age stratified, onetime cross-sectional serological survey in 2,184 adults and children from 25 communities were conducted in Beijing. The participants were investigated during April 15 to 18. This study had been approved by the IRB at Peking University Health Science Center prior to participant enrollment (IRB00001052-20021). Written informed consents were acquired from all participants in the investigation. For child <18 yrs, consent had been obtained from his/her legal guardian.

### Study population

Residents who aged > 1-year-old, lived in Beijing for at least 14 days in between of January and March 2020 were eligible for participation.

### Sampling strategies

This survey was performed by a multi-stage cluster random sampling technique. The highest four incidence districts (Xicheng, Shijingshan, Daxing and Fengtai) and the lowest incidence district (Pinggu with no confirmed case reported) out of 16 districts in Beijing were selected according to the protocol by World Health Organization [5]. Five communities in each district and certain households in each community were randomly selected based on Probability Proportionate to Size (PPS) at two stages respectively. A household was defined as a group of people (1 or more) living in the same residence. All persons living in the household had been invited to participate in the study, including children. If the age structure of samples were not consistent with that of general population in targeted communities due to refusal or contraindication to venipuncture, additional households were randomly selected to ensure the representativeness of samples.

### Data collection

We recruited participants organized by community residential committee. Each participant enrolled in survey was asked to fulfill a concise questionnaire which covers demographic information and COVID-19 relevant exposure information.

### Specimen collection

A certain amount of whole blood sample was drew into vacutainer without anticoagulant from each participant (5ml for 5+ years old, 3ml for the rest) and transported to the local Center for Disease Control and Prevention (CDC) laboratory. The serum was separated and apportioned into two aliquots A and B. The serum in tube A was transported to Beijing CDC laboratory for SARS-CoV-2 antibody testing within two days, while tube B was stored at -40°C for backup. All the procedures were according to COVID-19 laboratory guidelines in China.

### Serological testing

Serum samples were screened for the presence of SARS-CoV-2 specific antibodies using two different antibody test (Colloidal Gold) kits, Wondfo (Guangzhou, Batch W19500315, testing total antibody) and Innovita (Tangshan, Batch 20200402, testing IgM/IgG), which had been approved by China Food and Drug Administration (CFDA). Test kits were read in 15 minutes. According to the manufacturer, test performance characteristics showed a sensitivity of 86.4% (95% CI 82.5-89.6%) and a specificity of 99.6% (95% CI 97.6- 99.9%) for Wondfo and a sensitivity of 87.3% (95% CI 80.4-92.0%) and a specificity of 100% (95% CI 94.2-100%) for Innovita. Both test performance also verified on a sample of 20 PCR-confirmed COVID-19 positive patients and 10 gold-standard negative serum specimens collected before COVID epidemic. The sensitivity and specificity were 100% for both test kits. The testing procedures were carried out in Beijing CDC laboratory with biosafety level 2 (BSL-2) capacity.

If a serum sample was positive when using either test kit, the serum in tube B (backup serum) would be couriered to China CDC for a micro-neutralization assay to measure and confirm the SARS-CoV-2 -specific neutralizing antibody. For seronegativity in colloidal gold tests, 20 random samples were confirmed using the same micro-neutralization assay. Serum samples were inactivated at 56°C for 30 minutes and serially diluted with cell culture medium in two-fold steps. The diluted serums were mixed with a virus suspension of 100 TCID50 (50 tissue culture infective dose) in 96-well plates at a ratio of 1:1, followed by 2 hours incubation at 36.5°C in a 5% CO_2_ incubator. 1-2×10^4^ Vero cells were then added to the serum-virus mixture, and the plates were incubated for 5 days at 36.5°C in a 5% CO_2_ incubator. Cytopathic effect (CPE) of each well was recorded under microscopes, and the neutralizing titer was calculated by the dilution number of 50% protective condition. A titer of 1:4 or higher indicated seropositivity.

### Statistical analyses

Age-specific cumulative incidence was the proportion of participants per age strata who tested seropositive for SARS-CoV-2 infection. Proportion were adjusted for difference in the age structure of the participants and the overall population. Population weighting estimation and more details were shown in the Statistical Appendix.

We used a likelihood ratio test to calculate 95% confidence intervals of fractions with the Clopper-Pearson exact method (when the estimated fraction was 0), as implemented in the R package binom [3].

## Results

From 39,769 households in 5 districts of Beijing, 578 households, including 1,510 subjects in the sampling protocol participated in this study. Considering the age structure of 1,510 subjects were not consistent with that of the local populations, additional subjects were randomly selected in certain age groups. Between April 15 and 18, 2020, a total of 1,247 households including 2,184 participants were enrolled in this research.

Table 1 provided demographic characteristic of unadjusted sample and population-weight adjusted of the sample and Beijing estimates. The sample distribution was not significantly deviated from that of Beijing.

**Table 1.**
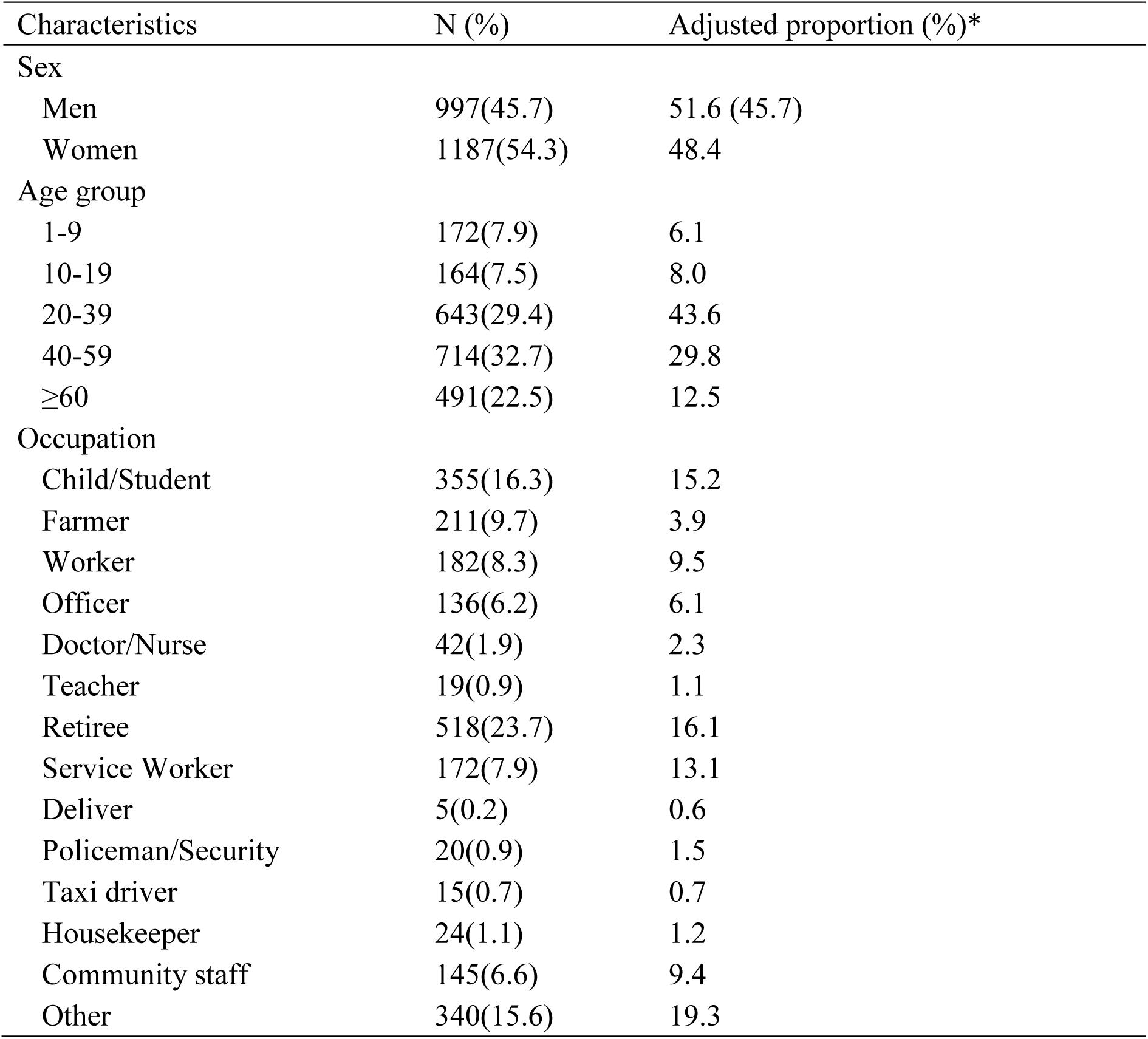
Demographic characteristics of 2,184 study participants

### SARS-CoV-2 infection

Thirteen of the 2,184 serum samples were tested seropositive by Colloidal Gold tests, among which 8 were IgM positive, 3 were IgG positive, 1 was total antibody positive, 1 was both IgM and total antibody positive.

None of the 13 colloidal gold positive individuals reported any possible exposure (Table 2).

**Table 2.** Serological results in 2,184 study participants.

| Variable | Total<br>(N=2184) | Colloidal gold test<br>Positive (N=13) | Micro-neutralization assay<br>Positive (N=0) |
| --- | --- | --- | --- |
| Male sex - no. (%) | 997(45.7) | 7(53.9) | - |
| Mean age - yr | 42.3±19.5 | 45.6±18.3 | - |
| Patients with chronic<br>Diseases - no. (%) | 541(24.8) | 1(7.7) |  |
| Pregnant<br>Women - no. (%) | 4(0.2) | 0(0) |  |
| Any travel – no. (%) | 63(2.9) | 0(0) | - |
| Specific travel – no. (%) |  |  |  |
| Wuhan | 3(0.1) | 0(0) | - |
| Hubei Province* (Wuhan<br>excluded) | 46(2.1) | 0(0) | - |
| International travel | 15(0.7) | 0(0) | - |
| Contact of patients with<br>symptom(s)** -no. (%) | 29(1.3) | 0(0) | - |
| Contact with infected cases – no.<br>(%) | 3(0.1) | 0(0) | - |
| Reported symptom(s)** – no. (%) | 75(3.4) | 0(0) | - |
| Reported any clinic visit history –<br>no. (%) | 53(2.4) | 0(0) | - |
| Reported being diagnosed of<br>COVID-19 | 0(0) | 0(0) | - |

\* Wuhan is the capital city of Hubei province.
\*\* All-caused respiratory symptoms including fever, cough, shortness of breath, or pneumonia, do not limit to COVID-19.

Among 13 seropositive samples and 50 randomly selected seronegative samples, none was positive in neutralization assay.

Considering no positive result in neutralization assay, we did not estimate the age-specific cumulative incidence and the adjusted seroprevalence.

Using a likelihood ratio method to calculate 95% confidence intervals of fractions with the Clopper-Pearson exact method, we estimated that the seroprevalence of COVID-19 was, with 95% confidence, not higher than 0.17% in Beijing.

## Discussion

In this study, we utilized three independent testing methods to investigate the seroprevalence of SARS-CoV- 2 antibody in a multi-stage cluster random sample including 2,184 participants in five districts of Beijing during April 15 to 18. No positive neutralizing antibodies case was detected by micro-neutralization assay.

We estimated that the percentage of participants that tested positive in population screening were not higher than 0.17% with 95% confidence during April 15-18, which indicates Beijing’s control effort had curtailed morbidity from COVID-19 and stopped community transmission of SARS-CoV-2 substantially. Meanwhile, these results also showed that the majority of populations in Beijing remained susceptible to SARS-CoV-2. Indeed, Beijing experienced a COVID-19 rebound in June. On June 11, a new COVID-19 case was reported in Beijing after no new confirmed infection for 56 consecutive days.

To date, micro-neutralization assay, as the gold-standard is the most specific and sensitive serological assay for evaluating and detecting functional neutralizing antibodies [6]. The updated seroeipdemiological investigation protocol of WHO (Version 2.0 released on 26 May) recommended that if a sample was positive or equivocal for either IgM, IgA or IgG using Rapid Diagnostic Tests (RDT), a neutralizing assay should ideally be performed [7]. Additionally, some studies showed that asymptomatic individuals had weaker immune response to SARS-CoV-2 infection comparing with symptomatic patients. The neutralizing antibodies of asymptomatic cases decreased within 2-3 months after infection [8]. Therefore, the seroprevalence might be underestimated if: (1) the serum sample was not collected within the window of neutralizing antibody production; (2) the immune response of asymptomatic infection was not strong enough to be detected.

Several research teams worldwide had started testing samples for SARS-CoV-2 antibodies. Among high risk populations, the average seroprevalence in healthcare a tertiary hospital in Germany in employees of a university hospital emergency department in USA was 1.6 % [9] and 5.9% [10] respectively. Among blood donors, 4.4-10.8% in Milan of Italy [11] and 5/500 in Scotland of UK [12] were positive for anti- SARS-CoV-2. Among general populations, 0.6% [3], 1.5% [2], 4.1% [4] and 22% [1] were antibody positive in random community participants in four surveys conducted in Iceland, Santa Clara and Los Angeles County of USA and Guilan of Iran respectively. One systematic review showed that the pooled percentage of asymptomatic infection was 46% (95% CI, 18.5-73.6%) [13]. The varying rates of COVID-19 antibody seropositivity in above studies emphasized the importance of serological survey in different populations. A study from the city of Wuhan, the epicenter of the COVID-19 pandemic in China, showed that the seropositivity here varied between 3.2% and 3.8% in non-random sampling of high risk populations including health workers, hotel staff members and family members of the healthcare workers [14]. More studies were needed to determine the seroprevalence in community residents in both high and low COVID- 19 epidemic regions of China.

Our study has several limitations. One of the main limitations is the performance of test kits. According to the manufacturer, the two test kits were both used for the additional testing for suspected case with negative PCR, which indicated that they are unsuitable for general population screening. Although none was positive in neutralization assay among these 13 seropositive samples and 20 randomly selected seronegative samples, we could not preclude the possibility of false negativity since the neutralization assay was conducted only in targeted part of residents. Despite the acceptable sensitivity of the two colloidal gold methods provided by the manufacturers, Döhla et al. found that antibody-based rapid test showed low sensitivity (36.4%) in high-prevalence community setting. They recommended not to rely on an antibody-based rapid test for public health measures such as community screenings [15]. Considering that the sample sizes of patients and controls in our validation were limited, we used test performance data in manufacturer instructions to establish the test’s sensitivity and specificity [2]. Additional validation of the assays especially in general populations used could improve further our estimates. The availability of other high-quality serological testing kits suitable for general population screening was expected. Inadequate sample size and one-time cross-sectional study performed were also needed to be considered. Given the low seroprevalence in Beijing, larger sample size is expected to be further investigated in order to achieve dynamic and more accurate seroprevalence estimation.

## Conclusions

The seroprevalence of COVID-19 was low in Beijing by April, which suggests that the comprehensive control measures effectively prevent and contain further spread in Beijing. However, the majority of the residents in Beijing were susceptible to infection and the risk of rebound should be noted due to the low population-level immunity.

## Data Availability

All data are available after the agreement of corresponding author.

## Acknowledgement

We thank China CDC for assistance with study design and laboratory support. The authors would like to thank all the investigators from Xicheng, Shijingshan, Daxing, Fengtai, and Pinggu CDC for field investigation and data collection and all participants for their contribution in this study.

